# Diagnostic-avoiding *Chlamydia trachomatis* variants detected in cervical and endometrial specimens from women during 16S microbiome profiling

**DOI:** 10.1101/2024.11.13.24316066

**Authors:** Sangmi Jeong, Tammy Tollison, Hayden Brochu, Hsuan Chou, Tammy Yu, Priyanka Baghaie, Kacy S. Yount, Toni Darville, Harold C Wiesenfeld, Sharon L Hillier, Xinxia Peng, Catherine M. O’Connell

**Affiliations:** North Carolina State University; University of North Carolina at Chapel Hill; University of Pittsburgh School of Medicine and the Magee-Womens Research Institute

**Keywords:** NAAT, chlamydia, gonorrhea, microbiome, 16S rRNA, false negative, 23S

## Abstract

**Background:** Performance of a 16S rRNA analysis of the cervicovaginal microbiome of 220 participants recruited into the T Cell Response against Chlamydia (TRAC) cohort between February 2011 and August 2014 in Allegheny County, Pennsylvania USA detected DNA encoding chlamydial 16S rRNA in samples from seven participants whose tests were negative for *Chlamydia trachomatis* (CT) and DNA encoding gonococcal 16S rRNA from five participants whose tests were negative for *Neisseria gonorrhoeae* (NG) infection with the Aptima Combo2 assay (Hologic).

**Methods:** We used targeted PCR amplification followed by sequencing to characterize the chlamydial 23S rRNA locus and qPCR to detect gonococcal DNA in residual diagnostic swab eluates or DNA used to generate 16S rRNA libraries.

**Results:** Discrepant specimens that contained chlamydial DNA carried a diagnostic-avoidant, G1526A variant in the 23S rRNA locus identical to variants previously detected in Finland, Denmark, and the UK. PCR validation of gonococcal DNA was confirmed for all participants whose tests were negative, with stochastic effects consistent with infection levels close to the limit of detection by the diagnostic assay.

**Conclusions:** These data indicate that this probe-avoidant CT mutant, and possibly others, were circulating in the northeastern US prior to their detection and characterization in 2019. Although infrequent, documentation of false negative results for CT indicates a need for clinicians to consider performance of a second test that uses alternate PCR targets if patients have persistent symptoms or have known contact to an infected sex partner and their initial NAAT is negative.

## Background

Performing cross sectional or longitudinal research studies investigating natural immunity or immunopathogenesis of sexually transmitted infections (STI) in humans can be a complex, time consuming and expensive undertaking. Efforts to mine all data associated with infection or disease cohorts are often extensive and protracted. Such is the case for the TRAC (T cell Response Against Chlamydia) cohort, a group of 246 cisgender women at elevated risk for chlamydial infection who enrolled between February 2011 and August 2014 while presenting to the Allegheny County, PA Health Department’s Sexually Transmitted Diseases Clinic for STI testing and/or management. The TRAC study sought to profile cell mediated responses associated with protection against ascending *C. trachomatis* (CT) infection (from the cervix to endometrium) or against CT reinfection [1, 2]. Since completion of enrollment, data obtained from participant samples and questionnaire responses have been used to profile antibody [1, 3–5], T cell antigen-specific responses [2], and to determine risk factors for ascending infection. Using modern molecular techniques, we quantified pathogen burden [1, 2], local cytokine secretion [6] and used genetic profiling techniques to search for biomarkers predictive of disease or reinfection risk [7–9]. Secondary investigations have also assessed the impact of *Mycoplasma genitalium* infection in this population [10]. Fundamental to the success of these analyses has been accurate classification of study participants according to infection status at enrollment and during subsequent follow up, so enrollee infection status was assigned according to the outcome of an FDA-cleared STI diagnostic NAAT test performed at the study visit, the results of which were used to inform clinical care. Infrequently, discrepant results were observed between research PCR assays detecting chlamydial loci and results obtained using the Aptima Combo 2® assay (AC2), manufactured by Hologic, Inc., a commercially available nucleic acid amplification test (NAAT) for CT and *Neisseria gonorrhoeae* (NG), targeting 23S ribosomal RNA (rRNA) and 16S rRNA in these bacteria, respectively. However, we could not exclude the possibility of cross-contamination from the environment or chlamydia-infected specimens, particularly with repurposed, reprocessed convenience samples, e.g., leftover transport medium.

The availability of pristine “future use” vaginal swabs collected from the same cohort and stored since enrollment provided an opportunity to reexamine these discrepant results while determining cervicovaginal microbiome profiles that associated with ascending chlamydial infection [11]. We performed a 16S rRNA gene genomic DNA based microbiome analysis on the entire cohort and detected chlamydial DNA in samples from seven participants whose tests were negative for CT and gonococcal DNA from five participants whose tests were negative for NG with the AC2 assay. PCR amplification and subsequent sequencing using primers directed against the 23S rRNA locus revealed that some of the specimens contained chlamydial DNA carrying a diagnostic-avoidant single nucleotide polymorphism (SNP) that likely contributed to misclassification. These data indicate that this mutant, and possibly others, were circulating in the northeastern US prior to their detection and characterization in the United Kingdom and Denmark in 2019 and subsequently [12–14]. Discrepant NG detection reflected very low gonococcal loads that challenged the limit of resolution for the NAAT. Although infrequent, documentation of false negative results for CT or NG may indicate a need for clinicians to consider performance of a second test that uses alternate or additional PCR probes.

## Methods

### T Cell Response against Chlamydia (TRAC) Cohort (246 participants)

This cohort was composed of young (median age, 21 years; range, 18-35 years) cis-gender women at high risk for the acquisition of CT [1]. Criteria indicating high-risk status included > 3 sexual partners in the previous 6 months, ≤ 14 years of age at sexual debut, history of pelvic inflammatory disease (PID), the presence of mucopurulent cervicitis on exam, or sexual contact with a partner known to be infected with CT or NG or non-gonococcal non-chlamydial urethritis. Women with a current diagnosis of PID were excluded. Additional exclusion criteria were pregnancy, uterine procedure or miscarriage in the preceding 60 days, menopause, hysterectomy, antibiotic therapy in the preceding 14 days, and allergy to study medications. TRAC participants were enrolled into a longitudinal study designed to investigate T cell responses important for protection from incident chlamydial infection over 12 months of follow-up between February 2011 and August 2014 and were recruited from the Allegheny County Health Department’s Sexually Transmitted Diseases Clinic, UPMC Magee-Womens Hospital (MWH) Ambulatory Care Clinic, and the Reproductive Infectious Disease Research Unit at MWH in Pittsburgh PA. Participants provided informed consent at the time of enrollment and completed questionnaires regarding obstetric/gynecologic history, behavioral practices, sex exposure, contraceptive methods, and symptoms. Clinical, histological, and microbiological testing was performed, and blood and endometrial samples were obtained after enrollment, after which all participants received single-dose agents for gonorrhea (ceftriaxone, 250 mg intramuscularly) and chlamydia (azithromycin, 1 g orally). Participants in this cohort were assessed for cervical and endometrial infection using AC2 (Hologic, Marlborough, MA) NAAT with overall infection rates of 67% and 8.5% for CT and NG, respectively. *M. genitalium* (MG) infection was determined using the Aptima MG NAAT (Hologic).

### 16S library construction and characterization

Cervicovaginal swab samples available from 246 women enrolled in TRAC were processed for 16S rRNA gene microbiome profiling [11]. In brief, cervicovaginal microbial communities present in participant specimens were profiled by sequencing the V4 region of 16S rRNA gene using Illumina sequencing. The 16S rRNA amplicon sequencing data was processed at the level of inferred amplicon sequence variants (ASVs) using USEARCH v11 [15]. The raw sequencing data supporting the conclusions of this article is publicly available at NCBI’s Short Read Archive (SRA) under BioProject accession number PRJNA1136868.

### PCR template purification from residual diagnostic samples

DNA was extracted as previously published using a Quick-DNA Universal Kit (Zymo Research, Irvine, California, USA) from 100 μL to 300 μL of reserved cervical swab eluates. Chlamydial and/or mycoplasmal loads were measured by quantitative PCR [1] targeting the 16SrRNA gene [16] or the highly conserved MG190 locus [10], respectively. Primers directed against the *parA* pseudogene [17], 16S rRNA and 23S rRNA loci [18] were similarly used to validate and/or quantify NG abundance.

### PCR amplification and sequencing of 23S rRNA alleles

Amplification conditions for detection of SNPs in *C. trachomatis* 23S were as described [19]. The melting temperature was raised to 98°C for optimal activation of the high fidelity, proof-reading enzyme Phusion (ThermoFisher). Samples were processed independently in a dedicated PCR hood to minimize risk of cross-contamination during analysis. Two rounds of amplification were required to recover sufficient product which was cleaned and concentrated using a DNA Clean and Concentrate kit (Zymo) before sequencing in forward and reverse directions using V3 region primers [19]. Two patient samples that had tested positively using the AC2 test were also analyzed as controls. Sequences generated via Sanger sequencing were aligned against the published sequence using Clone Manager Professional Version 11 (Sci Ed Software LLC, Westminster, CO). Residual cervical templates were used from profiled participants except for 3-209, for whom only endometrium-extracted DNA remained available for analysis.

### Statistical analysis

The nonparametric Spearman rank correlation coefficient was calculated to assess the overall correlation between CT 16S rRNA gene DNA copy numbers measured by qPCR and the raw read counts of CT 16S rRNA gene from microbiome sequencing analysis using R function ‘cor’ [20]. The LOESS (Locally Weighted Scatter-plot Smoother) curve was estimated and plotted using a ‘geom_smooth’ function of the R ‘ggplot2’ package with default settings [21]. Abundance of NG-specific ASVs present in libraries from individuals testing positively with AC2 and those who tested negative were compared using a Mann-Whitney test (two tailed).

## Results

### Detection of chlamydia-derived amplicon sequence variant (ASV) in 16S libraries derived from vaginal swabs obtained from CT negative TRAC participants

Total DNA was extracted from “future use” vaginal swabs from a total of 246 TRAC participants for inclusion in a 16S rRNA gene-based sequencing analysis of the contribution of the cervicovaginal microbiome to ascending chlamydial infection. Three samples were excluded based on DNA low integrity values after extraction. We also excluded twenty-three samples due to the equivocal results of chlamydial infection at the cervix and/or endometrium from clinical diagnostic testing. The remaining 220 samples were processed using the ASV-level analysis, resulting in 1,990 ASVs detected in total from samples sequenced to the average depth of ∼151,000 reads. Taxonomy annotations of ASVs were based on sequence comparison against the 16S rRNA gene reference database RDP version 19 [22] using the SINTAX algorithm [23]. We assessed the concordance of STI detection by examining raw read counts of sexually transmitted pathogen-specific 16S rRNA genes vs infections previously assessed by NAAT diagnostic tests (CT, NG, MG) (**Fig. 1a**) and quantified via qPCR (CT [1], **Fig. 1b**). About 58% (83 of 143) samples from participants whose tests were positive for CT at enrollment had non-zero 16S rRNA gene read counts (**Fig. 1a**). Similarly, 85% (17 of 20) samples from participants whose tests were postive for NG, and 15.8% (6 of 38) samples from MG positive enrollees had non-zero 16S rRNA gene read counts. Overall, there was modest correlation (Spearman = 0.575) between the two CT quantification methods (**Fig. 1b**).

**Figure 1.**
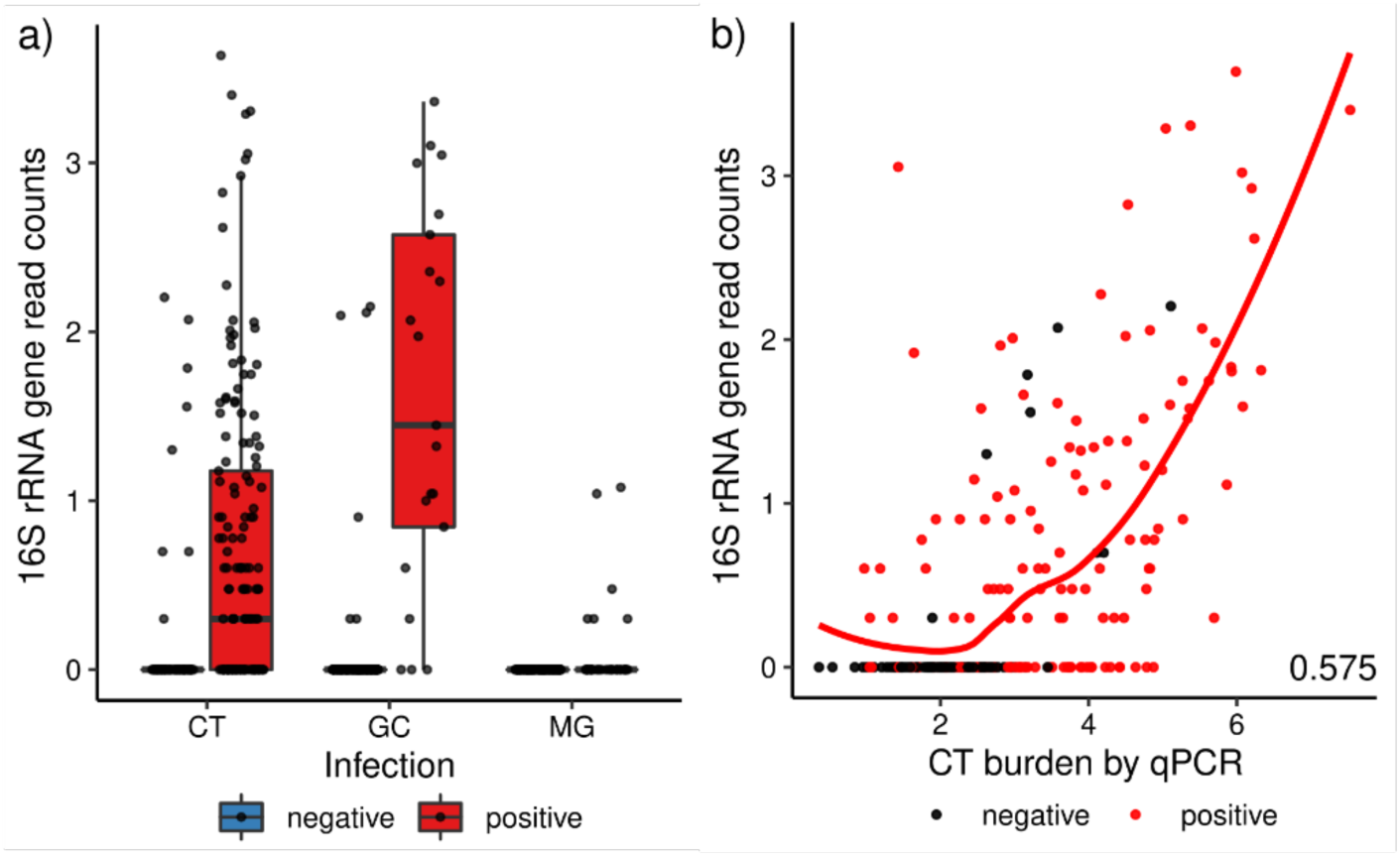
STI-derived 16S transcripts detected in vaginal swabs. a) 16S rRNA gene read counts of STIs stratified by infection status at enrollment. b) Concordance between CT 16S rRNA gene read counts versus CT burden by qPCR, where samples are colored by infection status at enrollment. A LOESS curve is shown in red and Spearman correlation in the bottom-right. CT, *Chlamydia trachomatis*; GC, *N. gonorrhoeae*; MG, *M. genitalium*

Based on clinical diagnostic results obtained at enrollment, the TRAC participants were classified into three groups: 1) Uninfected; 2) Endo-participants with CT infection detected only in their cervix; 3) Endo+ participants who tested positively for CT in cervix and endometrium. There were no TRAC enrollees whose tests were positive only in the endometrial sample. Comparing ASV abundances between CT-positive (Endo- and Endo+) and CT-negative (control) women, we were surprised to detect 16S ASV within libraries derived from samples from women who had been assigned “uninfected” (N=7) based on the results of NAAT testing (**Fig. 1a**). These results correlated with estimated chlamydial abundance derived from a previously performed quantitative 16S PCR assay (**Fig. 1b** and **Table 1**), suggesting that attributing the original results to external or cross-contamination was incorrect. A review of these participants’ clinical data revealed that they were mostly asymptomatic (5/7), one had reported experiencing dysuria, consistent with >10 leucocytes detected per high power field in her urine sample with irregular vaginal bleeding and another reported dyspareunia and abnormal vaginal discharge. Clinical findings upon pelvic examination were unremarkable for all. Three of the seven participants within this group reported contact with a CT infected partner in the three months prior to their enrollment but had yet to be treated.

**Table 1.** TRAC participants with discrepant clinical and molecular CT detection.

| Sample ID | Clinical testing (CT, MG, NG) | 16S rRNA read counts in ASV analysis | 16S qPCR* |  |
| --- | --- | --- | --- | --- |
|  |  |  | Cx | Endo |
| 3-101 | CT-/MG-/NG- | 1 | 78 | 919 |
| 3-144 |  | 16 | 420 | 87 |
| 3-145 |  | 146 | 128800 | 35 |
| 3-148 |  | 111 | 3839 | 457 |
| 3-201 |  | 55 | 1495 | 721 |
| 3-226 |  | 31 | 1640 | 105 |
| 3-209 | CT-/MG+/NG- | 4 | 13000 | 62680 |

\*Chlamydial 16S rDNA copies/swab [1].

In the context of the TRAC cohort, reasons for false negative CT NAAT test results, other than testing failures detected at or during performance e.g., compromised reagents or automated run failure (batch specific), include inadequate sampling at the anatomical site or an unanticipated flaw in the diagnostic test. All NAAT tests were run within temporal parameters appropriate for clinical STI testing, so batch-specific effects did not serve as an explanation for these discrepancies because the participants were not recruited contemporaneously. Testing of an endometrial biopsy might result in a false negative because the entire endometrial lining is not sampled, the amounts of tissue obtained may vary [7], and the region of specimen dedicated to NAAT testing is small [10]. However, cervical testing, particularly when performed by highly trained practitioners, is robust [24, 25]. Several publications from Northern Europe have reported instances of false negative testing specifically associated with the AC2 test [12, 13, 26, 27] arising from probe-avoidant mutations at the chlamydial 23S rRNA locus [19]. Speculating that this might explain our discrepant results, we attempted to amplify the relevant V3 region from cervically or endometrially derived templates that had been extracted from residual diagnostic swab eluates. In all instances we were successful using the protocol and primers used by Hokynar et al. [19], further confirming the presence of chlamydial DNA in the clinical samples. We detected a G-A substitution at 1523 in the samples of all seven test-discrepant participants (**Fig. 2**) when these products were sequenced. NAAT positively-testing samples (N=2) from the cohort were wild type at this position (data not shown). These data indicated that at least one probe-avoidant variant of CT was circulating in the region between May 2012 and February 2014, the time window between enrollments of our earliest and latest discrepant TRAC participants.

**Figure 2.**
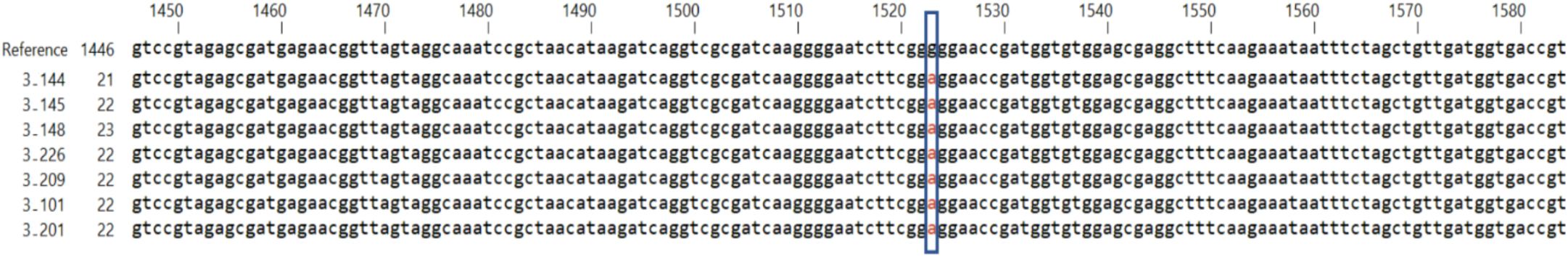
Portion of sequence alignment of V3 amplified PCR products from discrepant TRAC participant specimens revealing a g to a substitution at 1523 in the 23S rDNA locus. PCR products from two representative CT+ samples, as determined by AC2 diagnostic test were wild type at this position (data not shown).

16S rRNA gene microbiome analysis also revealed a total of five participants with discrepant NG AC2 testing enrolled from April 2011 to July 2013 (**Fig. 1A**, **Table 2**), including one participant, 3-209, who also tested discrepantly for CT (**Table 1**). One participant reported mild urinary symptoms with abnormal vaginal discharge and irregular bleeding and one other participant reported bleeding with intercourse. Clinical findings on pelvic examination were noteworthy for bleeding with cervical contact for four of five participants but were otherwise unremarkable. Only one of these participants reported contact with an infected partner within the preceding three months but had not received treatment.

**Table 2.** *Neisseria sp.* transcripts detected by 16S ASV analysis and genes detected by qPCR in diagnostic-discrepant or AC participants.

| Sample ID | Diagnostic test |  | 16S rRNA read counts |  |  |  | qPCR† |  |  |
| --- | --- | --- | --- | --- | --- | --- | --- | --- | --- |
|  | Cervix | Endo | ZOtu95:<br>N_gonorrhoeae | ZOtu141:<br>N_subflava | ZOtu264:<br>N_mucosa | ZOtu1566:<br>N_elongata | 16S | 23S | ParA |
| 3-121 | - | - | 130 | 0 | 0 | 0 | + | + | - |
| 3-140 | - | - | 123 | 0 | 0 | 0 | + | + | + |
| 3-146 | - | - | 106 | 0 | 0 | 0 | + | + | ND |
| 3-151 | - | - | 6 | 0 | 0 | 0 | + | - | + |
| 3-209 | - | - | 1 | 0 | 0 | 0 | - | + | ND |
| 3-012 | + | - | 471 | 0 | 0 | 0 | + | + | + |
| 3-026 | + | - | 10 | 0 | 0 | 0 | - | - | - |
| 3-052 | + | - | 10 | 0 | 0 | 0 | - | + | - |
| 3-089 | + | - | 24 | 0 | 0 | 0 | - | + | + |
| 3-091 | + | - | 6 | 0 | 0 | 0 | + | + | + |
| 3-112 | + | - | 1032 | 0 | 0 | 0 | + | + | + |
| 3-114 | + | - | 85 | 0 | 0 | 0 | - | - | - |
| 3-120 | + | - | 19 | 211 | 131 | 0 | - | + | - |
| 3-122 | + | - | 110 | 0 | 0 | 0 | + | + | + |
| 3-124 | + | - | 927 | 0 | 0 | 0 | - | + | - |
| 3-133 | + | - | 3 | 0 | 0 | 0 | - | - | - |
| 3-136 | + | - | 2 | 0 | 1 | 0 | + | + | - |
| 3-177 | + | - | 180 | 0 | 0 | 0 | + | + | + |
| 3-208 | + | - | 1151 | 0 | 0 | 0 | - | + | - |
| 3-212 | + | - | 9 | 0 | 0 | 0 | + | + | + |
| 3-223 | + | - | 206 | 0 | 0 | 0 | + | + | + |
| 3-235 | + | - | 349 | 0 | 0 | 0 | + | + | + |

†Assay performed with diagnostic swab eluate or vaginal swab template used for 16S rRNA microbiome analysis. ND not done because specimen was insufficient or unavailable.

The genus *Neisseria* includes a group of commensal species, *N. subflava*, *N. mucosa*, *N. elongata*, in addition to the important human pathogens, *N. gonorrhoeae* and *N. meningitidis* with the potential for genetic exchange between these naturally competent microorganisms [28] so we initially speculated that the ASV detected lacked discriminatory capacity. However, alignment of the 16S V4 sequence from *N. gonorrhoeae* with the 35 *Neisseria* species included in the RDP v18 database, revealed that ASV ZOtu95, detected in the discrepant samples, was highly specific, aligning with 100% identity between bases 535-787 of the 1544 nucleotide V4 region. This suggested that the 16S ASV detected in libraries generated from the vaginal swabs reflected true gonococcal infection. Next, we compared the number of ZOtu95 reads from discordant individuals with those detected in libraries generated from TRAC participants with concordant NG detection (Table 2) as a surrogate for pathogen load. The number of reads counted in NG+ samples ranged across four orders of magnitude but participant swab samples yielding libraries with ≤6 counts still tested positively via AC2 and no difference was detected between the groups using the non-parametric Mann-Whitney test (P=0.4116). Low levels of infection may have contributed to differential sampling of bacterial populations present at the cervix (diagnostic) and in the vagina (16S analysis). Quantifying NG by qPCR in residual template was difficult because of the minimal amount of cervical DNA remaining and the potential for high genetic variability between strains. Using primers that targeted multicopy (16S, 23S rRNA) and single copy (ParA pseudogene) loci, with genomic DNA extracted from *N. gonorrhoeae* strain FA19 as a positive control [29], we performed qPCR on diagnostic swab eluates or the 16S template that remained from discrepant NG+ participants (Table 2). Evaluating the performance of the primer pairs using material obtained from participants testing positively for NG infection, we observed that only 8 of the 17 samples consistently yielded PCR product for all 3 loci, while 6, were positive for 1-2. We were unsuccessful in amplifying material from the remaining 3 samples. Overall, successful amplification of the targets did not reflect abundance of 16S read counts in participants’ 16S rRNA libraries. For samples derived from participants who had tested negatively clinically but with ZOtu95 reads detected via 16S rRNA microbiome profiling, all tested positively for at least one multicopy locus.

With no source for cross-contamination available in the lab performing the 16S rRNA analysis, we also looked retrospectively at participant enrollment dates to exclude the remote possibility of cross- or environmental contamination [30] occurring during sample collection or NAAT testing. In only one instance did we observe enrollment of a discrepant NG- (3-121) and a confirmed clinically NG+ participant within the same 48-hour window (3-120). However, the presence of additional, neisserial species ASVs in the library generated from 3-120 only (Table 2), is inconsistent with contamination of the discrepant NG-sample by material derived from this diagnostic swab.

## Discussion

This study revealed that non-congruent CT infection outcomes that we detected between clinical diagnostics and 16S analysis for cervicovaginal samples collected from TRAC enrollees are best explained as resulting from false negative NAAT tests. The point mutation in 23S that we detected in discordant TRAC participants has been associated with reduced probe binding with the original AC2 assay. The G1523A variant was detected in the UK after clinical samples were screened for probe-avoiding mutants that might be in circulation [12] and in Denmark [13]. This is the first report of its detection in the United States although Katz et al., reported other 23S variants that mapped to A1518G or G1526A escaping detection in specimens collected in a five-week window, spanning late 2019 to early 2020 (2 of 401) [31]. Our retrospective analysis suggests that diagnostic-avoidant CT was already circulating in western Pennsylvania, USA by 2012 or possibly earlier. The first participants of the TRAC cohort enrolled in February 2011, but we did not identify diagnostically discordant enrollment samples collected prior to May 2012. Provided that this mutation does not impair chlamydial fitness, it is likely that it continues to circulate. The first 23S SNP variant escaping detection by the AC2 assay, FI-nvCT (C1515T), is still present in Finland at an estimated prevalence of 1.8% [32]. Fortunately, the deficit in the clinical test has been recognized and remediated by the manufacturer via incorporation of an additional chlamydial target [26] to improve reliability.

While discordancy between the clinical diagnostic assay for CT 23S RNA and detection of CT 16S rRNA, whether by qPCR or in the 16S rRNA libraries generated for microbiome analysis was ultimately elucidated by our identification of a genetic variant, the basis for the discrepancy of detection of NG 16S ASVs in participant samples from study participants who were diagnosed NG negative via the AC2 assay is not easily explained. The NG probe of the AC2 assay is directed against 16S rRNA and is both sensitive and highly specific [33–35]. Multiplex diagnostic tests have been reported to have high sensitivity and high concordance with standard nucleic acid amplification testing [36, 37] with a 4-in-1 test for CT, NG, *Trichomonas vaginalis*, and *M. genitalium* receiving recent clearance by the FDA [38]. False positive results for NG have been obtained arising from recombination between 16S loci of *N. meningitidis*, now recognized as a urogenital pathogen [39–42] and NG [43, 44]. The presence of multiple ribosomal RNA operons in this genus [45] elevates the risk for even rare recombination events to become fixed in the population. Should the inverse recombination event also occur, and NG acquire 16S copies from other neisserial species, this could lead to reduced sensitivity of currently available diagnostic tests, the consequences of which could negatively impact infection control and promote spread of diagnostic-avoidant mutants as was observed with Swedish nvCT [46, 47]. Identification of highly conserved, alternative targets for detection of NG has proven extremely challenging, so it is not surprising that we were unable to positively confirm the presence of genomic DNA in all the samples. However, we were also unable to amplify even multicopy targets from specimens collected from participants that tested positively by clinical NAAT, raising the possibility of low abundance bacteria that are asymmetrically distributed between the cervix and vaginal vault of discrepant enrollees.

Fortunately, study participants of TRAC were not harmed by failure to detect their CT and/or NG infection because all recipients were administered appropriate antibiotic therapy at enrollment regardless of their NAAT result. Appropriately, health care providers are moving from empiric syndromic treatment of patients at risk for STIs to pathogen-specific treatment guided by results of molecular tests [48]. Untreated CT and NG infections can lead to severe complications including PID, infertility, ectopic pregnancy and chronic pelvic pain in women. While NAATs are highly accurate, our study demonstrates that clinicians and epidemiologists should remain vigilant for potential CT or NG variants evading current approved NAATs such as detection of negative test results in a recently exposed individual, or a decline in overall rates of infection in a particular geographic location as occurred with Swedish nvCT [46, 47].

## Data Availability

The raw sequencing data supporting the conclusions of this article is publicly available at the NCBI Short Read Archive (SRA) under BioProject accession number PRJNA1136868.

https://www.ncbi.nlm.nih.gov/bioproject/PRJNA1136868/

## Declarations

### Ethics approval and consent to participate

The study protocol was approved by the institutional review boards for human research of the University of Pittsburgh (#PRO10010159) and the University of North Carolina (#13-3074). All participants provided written informed consent at the time of enrollment.

### Consent for publication

Not applicable.

### Availability of data and materials

The raw sequencing data supporting the conclusions of this article is publicly available at NCBI’s Short Read Archive (SRA) under BioProject accession number PRJNA1136868.

### Authors’ contributions

SJ, KSY, XP, and CMO designed research studies. HCW and SLH directed collection of participant samples and performance of diagnostic testing. TT, HC, TY and PB performed experiments. SJ, TT, HB, HC, and XP analyzed data. SJ, KSY, TD, XP, and CMO wrote the manuscript. All authors reviewed the manuscript.

## Acknowledgements

We thank the women who agreed to participate in this study; Ingrid Macio, Melinda Petrina, Carol Priest, Abi Jett, and Lorna Rabe for their efforts in the clinic and the microbiology laboratory; and the staff at the Allegheny County Health Department STD Clinic, for their efforts. We would also like to acknowledge the contribution of Dr. Ian Huntress, who sadly passed away during the performance of these studies.

